# ACE inhibitors, AT1 receptor blockers and COVID-19: clinical epidemiology evidences for a continuation of treatments. The ACER-COVID study

**DOI:** 10.1101/2020.04.28.20078071

**Authors:** Luc Dauchet, Marc Lambert, Victoria Gauthier, Julien Poissy, Karine Faure, Alain Facon, Cécile Yelnik, Sophie Panaget, Thierry Plagnieux, Florent Verfaillie, Daniel Mathieu, Patrick Goldstein, Aline Meirhaeghe, Philippe Amouyel, on behalf of the Lille COVID-19 study group

**Author notes:** This clinical study took place at the University Hospital of Lille (Centre Hospitalier et Universitaire de Lille, CHU Lille). **Corresponding author** Prof. Philippe AMOUYEL, MD, PhD, Univ. Lille, Inserm, CHU Lille, Institut Pasteur de Lille, U1167 - RID-AGE - Risk factors and molecular determinant s o f aging-related diseases, Institut Pasteur de Lille, 1 rue Pr. Calmette, F-59000, Lille, France, E mail, ORCID: 0000-0001-9088-234X.

## Abstract

**Aims:** The question of interactions between the renin angiotensin aldosterone system drugs and the incidence and prognosis of COVID-19 infection has been raised by the medical community. We hypothesised that if patients treated with ACE inhibitors (ACEI) or AT1 receptor blockers (ARB) were more prone to SARS-CoV2 infection and had a worse prognosis than untreated patients, the prevalence of consumption of these drugs would be higher in patients with COVID-19 compared to the general population.

**Methods and results:** We used a clinical epidemiology approach based on the estimation of standardised prevalence ratio (SPR) of consumption of ACEI and ARB in four groups of patients (including 187 COVID-19 positive) with increasing severity referred to the University hospital of Lille and in three French reference samples (the exhaustive North population (n=1,569,968), a representative sample of the French population (n=414,046), a random sample of Lille area (n=1,584)).

The SPRs of ACEI and ARB did not differ as the severity of the COVID-19 patients increased, being similar to the regular consumption of these drugs in the North of France population with the same non-significant increase for both treatment (1.17 [0.83–1.67]). A statistically significant increase in the SPR of ARB (1.56 [1.02–2.39]) was observed in intensive care unit patients only. After stratification on obesity, this increase was limited to the high risk subgroup of obese patients.

**Conclusions:** Our results strongly support the recommendation that ACEI and ARB should be continued in the population and in COVID-19 positive patients, reinforcing the position of several scientific societies.

## Introduction

Since December 2019, the world is being facing the largest pandemic ever in this century. Although a large number of the COVID-19 patients will be asymptomatic or develop a mild disease, 5% (1) will be admitted to an ICU with an acute respiratory disease syndrome (ARDS) of highly severe prognostic. In a retrospective study, men aged over 68 years with hypertension and other cardiovascular comorbidities were more frequent among deceased patients (2). This observation of the association of cardiovascular comorbidities with severe COVID19 prognostic has been constantly reported in recent literature (3) where old age, cardiovascular disease, hypertension and diabetes were found to be prognostic factors for the COVID-19 related death. These cardiovascular conditions strongly benefit from the use of ACE inhibitors (ACEI) and AT1 receptor blockers (ARB) known to protect against hypertension, myocardial fibrosis and hypertrophy (4). These treatments became a large area of concern because of the key role of the Renin-Angiotensin-Aldosterone System ( RAAS) in the invasion of the SARS-CoV2 virus in human organisms (5). Indeed S ARS-CoV2 is known to target the host protein angiotensin converting enzyme 2 (ACE2) as a co-receptor to penetrate the intracellular compartment of human alveolar epithelial cells and other tissues (6). The ACE2, a homolog of ACE (7), is a key player of the beneficial effects of ACEI and ARB. ACE2 efficiently degrades angiotensin II, a powerful vasoconstrictor, to Ang (1–7) with anti-inflammatory and anti-fibrotic actions. Experimental studies tended to demonstrate that RAAS blockade enhances ACE2 properties by stimulating ACE2 expression and/or activity, contributing to the expected benefits of these drugs (8). These evidences prompted several questions: does this potential increase in the number of the molecular gates of the virus induced by RAAS blockers favours its dissemination? Should RAAS blockers be withdrawn in high risk patients with COVID-19 infection? Would this withdrawal modify the severity of the infection? Different reviews (5, 9, 10) have attempted to address these questions compiling what is known in the field of basic research, experimental medicine and clinical trials, with no definite conclusions, except that the lack of clear evidence should refrain physicians to withdraw these treatments.

To address this key issue, we used clinical epidemiology to estimate the prevalence of consumption of ACEI and ARB in COVID patients compared to the general population. We assumed that if an increased susceptibility to SARS-COV2 infection and/or a deleterious effect occurred in patients using ACEI or ARB, the higher the consumption of these treatments, the higher the risk of infection and the severity of the clinical symptoms. We tested this hypothesis in four groups of patients of different severity referred to the University hospital of Lille (North of France) compared to three reference samples.

## Methods

### Study oversight

This mono-centric study took place at the University Hospital of Lille (Centre Hospitalier et Universitaire de Lille, CHU Lille) and designed by the investigators, trained physicians and epidemiologists, in the context of the French reference methodology MR004, as a descriptive noninvasive epidemiological studies in human. All the data collected were compliant with the GDPR of the University Hospital. The investigators were never in contact with the patients or their families. The study was approved by the institutional review board of the CHU Lille and declared to the Commission National Informatique et Liberté.

### Patient samples and data collection

We collected clinical data from consecutive patients aged 35 years and over, referred to the CHU Lille for COVID-19 suspicion. We identified four groups of patients of different disease severity. In the first group, we registered all the individuals who called the national medical emergency number from 18^th^ March to 5^th^ April. Since the COVID-19 pandemic, each individual calling this number with any respiratory problems and/or cough and/or fever was immediately transferred to a dedicated COVID-19 call platform. A questionnaire was filled in to collect information on the current symptoms and the on-going anti-hypertensive treatments if any. The second group was composed of patients referred to an outpatient clinic dedicated to COVID-19 from 29^th^ February to 23^rd^ 26 March and benefited of genomic tests and clinical examination. The third group was composed of more severe COVID-19 positive cases who needed hospitalisation in a specific ward from 26^th^ February to 5^th^ April. The fourth group was composed of the most severe confirmed cases who needed to be hospitalised in intensive care units (ICU) for oxygenotherapy or mechanical ventilation from 21^st^ February to 3^rd^ April. Epidemiological, clinical and laboratory information, as well as risk factor levels and treatments at entry were extracted from the electronic medical records.

### Reference populations and data collection

We used three reference population samples to estimate the standardised prevalence ratio of anti-hypertensive treatment consumption. The first one was composed of the 2019 medical records of the exhaustive population of the North Department of France over 35 years of age (n=1,569,968) where the CHU Lille is located. The relevant data were extracted from the national health insurance information system (Système National d’Information Interrégimes de l’Assurance Maladie, SNIIRAM) created in 1999 in order to determine and evaluate more precisely health care utilisation and health care expenditure of beneficiaries. These nationwide data, based on almost 66 million inhabitants comprises individual information on the socio-demographic and medical characteristics of beneficiaries and all hospital care and office medicine reimbursements, coded according to various systems (11). The second one was a 1/97^th^ random sample of the SNIIRAM (Echantillon généraliste des bénéficiaires, EGB), representative of France’s national population of health insurance beneficiaries over 35 years of age in 2017 (n=414,046). For these two reference samples we extracted the numbers of beneficiaries that had either an ACEI, an ARB or any antihypertensive drug estimated by the presence of three or more reimbursements recorded of these products during the year (2019 for the North Department, 2017 for the global French population sample). Medicinal products were identified by ATC class (Anatomical Therapeutic Chemical Classification System). These numbers were calculated for each 5 year-age classes from 35 years old and each gender, estimating the expected number of individuals in each class of age and gender (12) that were prescribed ACEI, ARB and other antihypertensive drugs.

The third one was the 2005–2007 MONA LISA cross-sectional survey of cardiovascular risk factors and nutrition carried out by the French Lille MONICA (monitoring of trends and determinants in cardiovascular disease) centre (13). This sample included 1,584 men and women aged 35–75 living the Lille urban area, randomly selected from the electoral rolls. Detailed information on anthropometric, demographic and health data including treatments coded according to the ATC class were available. As for the two previous reference samples the numbers of individuals that had an ACEI, an ARB or any antihypertensive drug were calculated for each 5 year-age classes and each gender. In addition, a third stratification level was considered based on the Body Mass Index (BMI) coded in three classes as normal (18.5–25 kg/m^2^), overweight (25–30 kg/m^2^) and obese (≥30 kg/m^2^)

### Statistical analysis

As a continuous variable, age was expressed as mean and standard deviation. Categorical data were summarised as counts and percentages. For the 95% confidence interval of proportion where the numbers were less than 5, fluctuation interval was calculated according to binomial law. For each of the four groups of severity a standardised prevalence ratio of consumption of each type of drug and a 95% confidence interval were calculated (14). Firstly, an observed number of prescription was calculated; secondly, the number of consumption estimated was calculated by applying the straturm-specific prevalence rates by 5 year age classes and gender obtained from two reference populations: the exhaustive population of the North of France (R1) and the representative sample of the French population (R2). A third type of standardised prevalence ratio of consumption were calculated for the ICU sample stratified on age classes, gender and BMI classes obtained from the MONA LISA cross-sectional survey. We calculated the necessary number of individuals to include in our study based on comparison of two binomial proportions. Statistical analyses were performed using R software (version 3.3.1) (RCore Team, 2016) and Epitools package (version 0.5–7).

## Results

The total number of patients aged over 35 years suspected of or diagnosed with COVID-19 in the CHU Lille during the study period was 288 (Table 1). Among these, 62% were male and almost 70% ICU. COVID-19 was confirmed in 187 patients. An anti-hypertensive treatment was prescribed in 19.7% of the patients referred to the outpatient clinic, while for patients hospitalised in ICU who were all COVID-19 positive, this percentage was over fifty. The prevalence of diabetes almost doubled between patients in outpatient clinic and hospitalised patients and still doubled between hospitalised patients and patients transferred in ICU. No difference in respiratory disease prevalence could be observed, while chronic kidney and cardiovascular diseases prevalences were higher in hospitalised and ICU patients. The distribution of these comorbidities tends to highlight a different severity of the disease among the three groups. The emergency calls group of suspected COVID-19 individuals was very similar to the North of France population group and to the French National sample in term of gender distribution and prevalence of anti-hypertensive treatments.

**Table 1.**
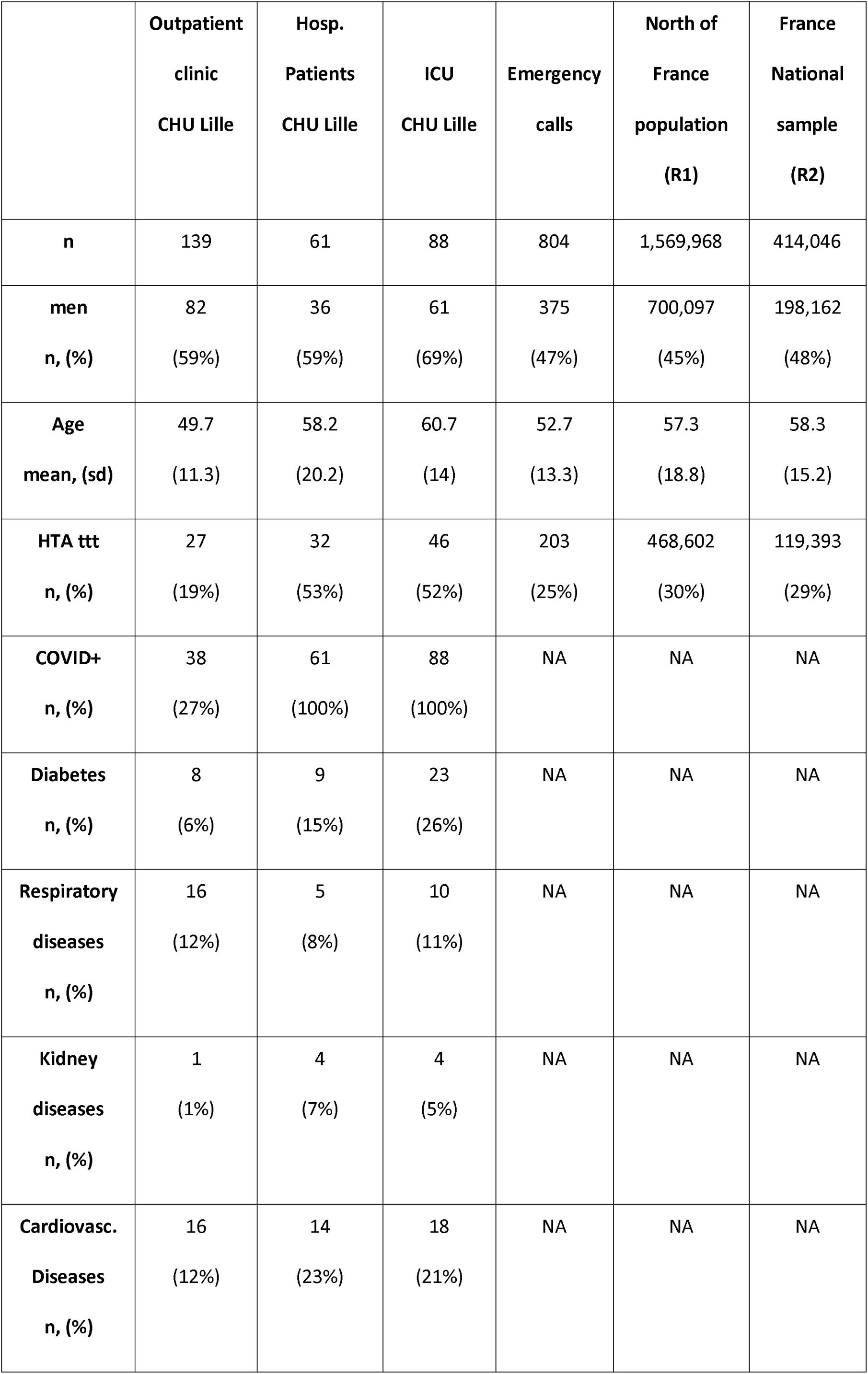
Description of the population samples.

In table 2, we calculated the prevalence of consumption of any anti-hypertensive drug, ACEI and ARB in the different groups of patients. More than half of the most severe cases, in hospital and ICU, were treated with antihypertensive drugs, this observation being consistent with the higher mean ages of these two samples and with hypertension as a potential prognostic factor for the COVID-19 severity. In a first attempt to estimate a possible additional increased risk of infection by SARS-CoV2 among patients treated by anti-hypertensive drugs, especially those treated with ACEI or ARB, we compared the proportion of ACEI and ARB use among all classes of anti-hypertensive treatments. These proportions were highly comparable (between 70 and 74%) between the two population reference samples and the one of the hospitalised and ICU patients, suggesting no clinically significant increase in ACEI and ARB prescription at entry, even for the more severe cases.

**Table 2.** Distribution of anti-hypertensive treatments.

|  | **N** | **Any anti**<br>**HTA drug**<br>**n (%)** | **ACEI**<br>**n (%)** | **ARB**<br>**n (%)** | **ACEI or ARB /**<br>**Any anti HTA**<br>**drug**<br>**%** |
| --- | --- | --- | --- | --- | --- |
| **Outpatient clinic**<br>**COVID +** | 38 | 7<br>(18%) | 4<br>(11%) | 2<br>(5%) | 86<br>[57-100]* |
| **Outpatient clinic**<br>**COVID -** | 101 | 20<br>(20%) | 8<br>(8%) | 9<br>(9%) | 85<br>[70-100]* |
| **Hospitalised**<br>**COVID+** | 61 | 32<br>(53%) | 14<br>(23%) | 8<br>(13%) | 69<br>[53-85] |
| **ICU**<br>**COVID +** | 88 | 46<br>(52%) | 13<br>(15%) | 21<br>(24%) | 74<br>[61-87] |
| **Emergency calls** | 804 | 203<br>(25%) | 70<br>(9%) | 82<br>(10%) | 74<sup>+</sup><br>[68-80] |
| **North of France**<br>**population (R1)** | 1,569,968 | 468,602<br>(30%) | 163,526<br>(10%) | 174,204<br>(11%) | 71<sup>+</sup><br>[70-71] |
| **France National**<br>**sample (R2)** | 414,046 | 119,393<br>(29%) | 38,760<br>(9%) | 46,931<br>(11%) | 70<sup>+</sup><br>[69-70] |

\* Fluctuation interval binomial law
<sup>+</sup> Individuals with association of ACEI and ARB were counted once only

When the reference population was the North of France population (Table 3), there was no significant difference between the standardised prevalence ratio (SPR) of the ACEI and ARB drugs in the COVID-19 positive patients. This absence of significant difference between the SPR of the ACEI and ARB drugs was also observed for all the four groups, except for patients in ICU where SPR for ARB drugs was significantly higher (1.56 [1.02–2.39]). This difference was in line with the other SPRs estimated with France National sample (1.63 [1.07–2.51]). To exclude a potential confounding factor in this increase in the use of ARB in ICU, conversely to what is observed in COVID-19 positive patients in outpatient clinic and hospitalisation, we stratified the ICU population according to obesity, a major prognostic factor of severity (15). In table 4, we calculated the SPR of the ACEI, ARB and other antihypertensive drugs using the MONA LISA Lille study, a representative sample of the Lille urban area as the reference group. We stratified the ICU population in three categories of BMI. In this severe prognostic sample, 43% of the patients were obese and 43% overweight. In ICU, the SPR increase in ARB, although not significant due to a lack of statistical power, was limited to the obese group. This suggested that the higher consumption of this drug may be related to the importance of the co-morbidity associated with obesity and not to a potential deleterious interaction with SARS-CoV2 infection. Accordingly, the median BMI in the obese patients in ICU (36.9) was higher than the one in the obese subjects of the MONA LISA Lille study (33.0).

**Table 3.** Standardised prevalence ratio of consumption of anti-hypertensive drugs.

|  | Treatment | Obs. | Exp. R1 | SPR1 [95%CI] | Exp. R2 | SPR2 [95%CI] |
| --- | --- | --- | --- | --- | --- | --- |
| **All patients** |  |  |  |  |  |  |
|  | ACEI | 31 | 26.4 | 1.17 [0.83-1.67] | 22.1 | 1.40 [0.99-1.99] |
| COVID + | ARB | 31 | 26.4 | 1.17 [0.83-1.67] | 25.4 | 1.22 [0.86-1.73] |
|  | Other drugs | 23 | 18.6 | 1.23 [0.82-1.86] | 17.7 | 1.30 [0.86-1.95] |
|  | ACEI | 78 | 76.1 | 1.03 [0.82-1.28] | 61.7 | 1.26 [1.01-1.58] |
| COVID - | ARB | 91 | 76.5 | 1.19 [0.97-1.46] | 72.2 | 1.26 [1.03-1.55] |
|  | Other drugs | 56 | 62.5 | 0.90 [0.69-1.17] | 57.2 | 0.98 [0.75-1.27] |
| **Emerg. calls** |  |  |  |  |  |  |
|  | ACEI | 70 | 68.8 | 1.02 [0.81-1.29] | 55.9 | 1.25 [0.99-1.58] |
|  | ARB | 82 | 69.7 | 1.18 [0.95-1.46] | 65.9 | 1.24 [1.00-1.55] |
|  | Other drugs | 53 | 57.0 | 0.93 [0.71-1.22] | 52.4 | 1.01 [0.77-1.32] |

| Outpatients |  |  |  |  |  |  |
| --- | --- | --- | --- | --- | --- | --- |
|  | ACEI | 4 | 3.2 | 1.24 [0.47-3.31] | 2.5 | 1.57 [0.59-4.19] |
| COVID + | ARB | 2 | 2.9 | 0.70 [0.18-2.80] | 2.7 | 0.74 [0.18-2.95] |
|  | Other drugs | 1 | 2.3 | 0.43 [0.06-3.07] | 2.0 | 0.50 [0.07-3.53] |
|  | ACEI | 8 | 7.3 | 1.10 [0.55-2.19] | 5.8 | 1.39 [0.69-2.78] |
| COVID - | ARB | 9 | 6.8 | 1.32 [0.69-2.54] | 6.3 | 1.42 [0.74-2.73] |
|  | Other drugs | 3 | 5.5 | 0.55 [0.18-1.70] | 4.8 | 0.62 [0.20-1.93] |
| Hospitalised |  |  |  |  |  |  |
|  | ACEI | 14 | 9.5 | 1.47 [0.87-2.49] | 8.1 | 1.72 [1.02-2.90] |
| COVID + | ARB | 8 | 10.1 | 0.79 [0.40-1.59] | 9.8 | 0.81 [0.41-1.63] |
|  | Other drugs | 10 | 7.1 | 1.41 [0.76-2.63] | 6.9 | 1.44 [0.78-2.68] |
| ICU |  |  |  |  |  |  |
|  | ACEI | 13 | 13.7 | 0.95 [0.55-1.64] | 11.4 | 1.14 [0.66-1.96] |
| COVID + | ARB | 21 | 13.5 | 1.56 [1.02-2.39] | 12.9 | 1.63 [1.07-2.51] |
|  | Other drugs | 12 | 9.3 | 1.30 [0.74-2.28] | 8.8 | 1.37 [0.78-2.41] |

2 R1 : North of France population reference, R2 : France National sample reference

**Table 4.** Distribution of anti-hypertensive drugs in ICU according to BMI.

|  |  | ACEI |  |  | ARB |  |  | Other drugs |  |  |
| --- | --- | --- | --- | --- | --- | --- | --- | --- | --- | --- |
|  | n | Obs. | Exp. | SPR<br>[95%CI] | Obs. | Exp. | SPR<br>[95%CI] | Obs. | Exp. | SPR<br>[95%CI] |
| **All** | 88 | 13 | 10.7 | 1.22<br>[0.71-2.09] | 21 | 15.5 | 1.36<br>[0.88-2.08] | 12 | 6.7 | 1.79<br>[1.02-3.15] |
| **Normal** | 12 | 1 | 1.2 | 0.86<br>[0.12-6.09] | 1 | 0.9 | 1.11<br>[0.16-7.91] | 4 | 0.7 | 5.58<br>[2.09-14.86] |
| **Overweight** | 38 | 5 | 5.4 | 0.93<br>[0.39-2.22] | 7 | 7.0 | 1<br>[0.47-2.09] | 3 | 2.6 | 1.14<br>[0.37-3.53] |
| **Obese** | 38 | 7 | 4.8 | 1.47<br>[0.70-3.08] | 13 | 9.4 | 1.38<br>[0.80-2.38] | 5 | 3.8 | 1.32<br>[0.55-3.16] |

4 Data calculated with the MONA LISA Lille study as the reference population

## Discussion

In this monocentric study, the standardised prevalence ratios of consumption of ACEI and ARB drugs were similar to the regular consumption of this drug in the North of France population and more generally, in a representative sample of France. In addition, these standardised prevalence ratios did not differ as the severity of the COVID-19 patients increased. However a statistically significant increase in the SPR of ARB drugs was observed in ICU only. After stratification on BMI, this increase of prescription was only observed in the obese patient group. This suggested a specific anti-hypertensive excess prescription of ARB in obese patients associated to a higher frequency of comorbidities, as hypertension and diabetes, and a highly deleterious prognostic for SARS-CoV2 infection in this subgroup of patients.

We defined four groups of suspected and confirmed COVID-19 cases with an increasing severity of their clinical form. If a deleterious relationship between the use of ACEI or ARB had existed, we would have expected an enrichment of patients consuming these drugs in severe cases. One limit of our study was the size of the COVID-19 positive sample. However, at least a doubling of the proportion of treatments could justify a strong clinical reason to eventually change the regular anti-hypertensive treatment use. The number of patients needed to detect a doubling of the prescription of ARB or ACEI (10% in the North of France reference sample) was 122 for α = 5% (2-tails) and β= 10%. When we grouped all the COVID-19 positive cases (187 cases), we did not detect any significant difference of prescription of ACEI or ARB.

When we estimated the standardised prevalence ratio in each of the groups according to the increasing severity, we observed a significant difference in the ICU patients compared to the two reference population samples, 21 patients treated with ARB were observed versus 13.5 and 12.9 expected, whereas these two numbers were similar among ACEI prescription. This could have been consistent with some basic evidences suggesting that the administration of ARB increased the level of cardiac ACE2 mRNA and activity while ACEI increased ACE2 mRNA levels only (16), ARB increasing the number of molecular gates for the SARS-CoV2. To better decipher this significant association, we explored in the ICU patients some of the risk factors of severity known to be associated with COVID-19. In particular, hypertension, estimated by the prevalence of use of anti-hypertensive drugs, had a frequency of 52% not different from the one observed in hospitalised patients (53%) where the use of ARB was not different from the one of the reference population samples (SPR1 = 0.79 [0.40–1.59]; SPR2 = 0.81 [0.41–1.63]). In these conditions, we could consider that this increase was not related to hypertension. Conversely, the frequency of diabetes was different between the different COVID-19 positive groups: 15% for the hospitalised patients and 26% for ICU patients. This prompted us to stratify our population sample in ICU according to a major risk factor associated with diabetes, increased cardiovascular risk and severe acute respiratory syndrome needing mechanical ventilation, i.e. obesity (15). Indeed, 43.2% of the ICU patients were obese and 43.2% were overweight. We stratified our samples according to age, gender and BMI in three classes and standardised our prevalence ratio of consumption with the expected number of subjects from of a random sample of the population were the ICU patients were located. The number of obese patients with ARB was 13 and the expected number was 9.4, while for overweight and normal patients these numbers were strictly identical. This result suggested that the doubling of the prescription of ARB in ICU patients was explained by the higher frequency of obese patients doubled in ICU (43.2%) compared to the one of the surrounding population (22.4%) (13). Obesity as a major prognosis risk factor for COVID-19 deserves further attention. Our European results are complementary to a retrospective study among hospitalised COVID-19 patients in China with hypertension where the inpatient use of ACEI/ARB was associated with a lower risk of all-cause mortality compared with ACEI/ARB non-users (17). In this study, the prescribing habits for hypertension were very different from the ones of the European populations. Indeed only 17% (188/1128) of their hypertensive patients were treated with ACEI/ARB while this figure was between 70 and 74% in our population samples. Therefore our results reinforced the interest to maintain ARB/ACEI treatments for COVID-19 positive patients in a European context of drug prescription. In addition to this Chinese sample, our study, taking into account non-hospitalised patients, supported the hypothesis that ACEI/ARB may not increase the probability of infection by SARS-CoV2 in the general population people.

In conclusion, our results suggest that the risks of infection by SARS-CoV2 and of an increased severity in patients suspected of or diagnosed with COVID-19 were not associated with an excess in consumption of ACEI and ARB (Summarising figure). Thus, based on the basic, experimental and clinical evidences of the high organ protective effects of ACEI and ARB, and despite all the theoretical deleterious hypotheses induced by the knowledge that the penetration gate of the SARS-CoV2 is the ACE2, our results strongly support the recommendation that RAAS inhibitors should be continued. Our results reinforce the position of several scientific societies (18,19). The results of our clinical epidemiology approach of the key question of the RAAS inhibitor in COVID-19 patients should be confirmed in other sample populations and countries around the world.

## Data Availability

The data can be obtained by contacting the corresponding author

## Acknowledgments

We thanks all the persons composing the Lille COVID-19 study group who have made substantial contributions to this manuscript (e.g. data acquisition (DA), analysis (A), or writing / editing assistance (E)) and helped to finalized this study in the shortest delays: Loic André^1^ (DA), Ady Assaf^2^ (DA), Edgar Bakhache^1^ (DA), Marcellin Bellonet^3^ (DA), Nordine Benameur^4^ (DA), Antoine Bidault^4^ (DA), Brigitte Bonneau^5^ (DA), Amélie Cerf^6^ (DA), Marie-Charlotte Chopin^2^ (DA), Brigitte Courtois^3^ (DA), Maureen Decroo^3^ (DA), Claire Delcourte^6^ (DA), Anne-Laure Demarty^3^ (DA), Dominique Deplanque^3^ (DA), Aurélie Dozier^2^ (DA), Marie-Pierre Dumont^7^ (DA), Emmanuel Faure^2^ (DA), Raphael Favory^6,7^ (DA), Claude Gady-Cherrier^8^ (DA and A), Corinne Gower^5,9^ (DA), Nathalie Guillon^5^ (DA), Mercé Jourdain^6^ (DA), Marine Letrionnaire^8^ (DA and A), David Messing^8^ (DA and A), Saad Nseir^6^ (DA), Hélène Pennel^5^ (DA), Anne Pétillon^5^ (DA), Mohammad-Ryad Pokeerbux^1^ (DA), Sébastien Préau^6,7^ (DA), Armand Rémy^4^ (DA), Renan Targhetta^3^ (DA), Sébastien Sanges^1^ (DA), Sarah Stabler^2^ (DA), Sophie Tallon^3^ (DA), Corinne Vantourout^5^ (DA), Fanny Vuotto^2^ (DA). (1. CHU Lille, Internal Medicine Department, F-59000, Lille, France; 2. CHU Lille, Department of Infectious Diseases, F-59000, Lille, France; 3. CHU Lille, Department of Research and Innovation, F-59000, Lille, France; 4. CHU Lille, SAMU 59 and Emergency Department, F-59000, Lille, France; 5. CHU Lille, Department of Public Health, Epidemiology, Medical Economy and Prevention, F-59000, Lille, France; 6. CHU Lille, Department of Intensive Care, F-59000, Lille, France; 7. Univ. Lille, Inserm, CHU Lille, Institut Pasteur de Lille, U1167 - RID-AGE - Risk factors and molecular determinants of aging-related diseases, F-59000, Lille, France; 8. DRSM Hauts de France, F-59009, Villeneuve d’Ascq, France; 9. Univ. Lille, Inserm, CHU Lille, U1286 – Infinite-Institute for Translational Research in Inflammation, F-59000, Lille, France). We are grateful to the medical students and the staff of the emergency call platform of the CHU Lille, to all the clinicians, the nurses and the non-medical staff, the Lille logistic departments and administrations of the CHU Lille involved in the COVID-19 care without whom this work would not have been possible.

## Take-home figure legend

No excess of consumption of ACEI and ARB as severity increased in patients with COVID-19 suggesting RAAS inhibitors should be continued. No more excess of consumption of ARB in ICU when stratified on Body Mass Index. (ACEI = Angiotensin Converting Enzyme Inhibitor; ARB = AT1 receptor blocker; ICU = Intensive Care Unit.)

## One-Sentence Summary

These results strongly support the recommendation to continue angiotensin converting enzyme inhibitors and angiotensin II receptor blockers in COVID-19 positive patients.

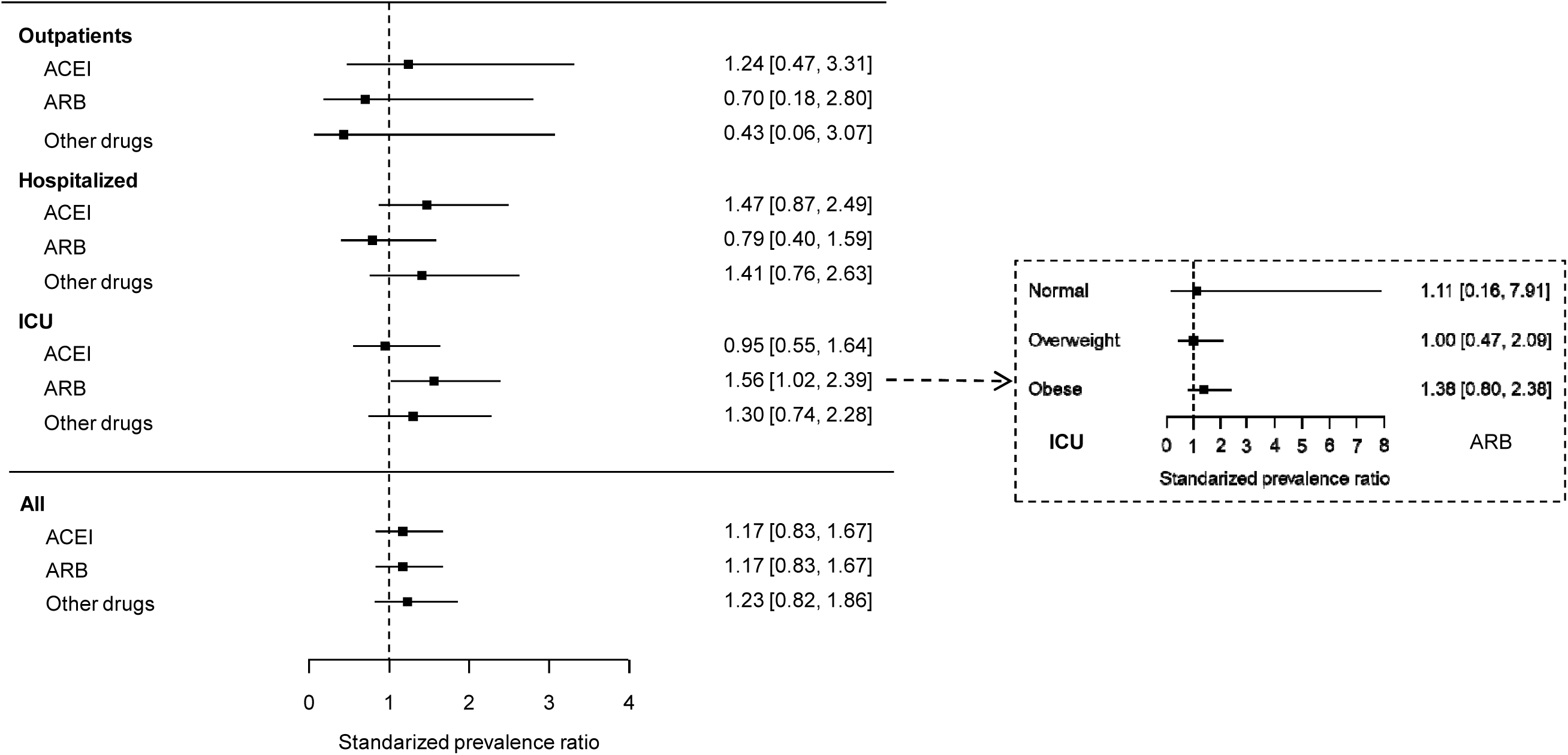

